# Association between thyroid function and thyroid homeostasis parameters and female urinary incontinence: A population-based study

**DOI:** 10.64898/2026.08.04.26359698

**Authors:** Junchao Zhang, Qian Yang, Jinfa Huang, Huan Yang, Ruolan Li, Kirtchhoof Tevorn, Kaixian Deng

**Author notes:** Department of Gynecology, The Eighth Affiliated Hospital, Southern Medical University (The First People’s Hospital of Shunde, Foshan), Foshan, Guangdong, China. Electronic address.

## Abstract

**Background:** Female urinary incontinence (UI) is a prevalent disorder associated with pelvic floor dysfunction. The abnormalities in neuromuscular function that cause the clinical symptoms of UI are rarely thought to be related to metabolic function, and may be especially related to thyroid hormone levels.

**Objective:** To examine the connection among thyroid function, thyroid homeostasis parameters, and UI, along with its various subtypes in females.

**Methods:** A total of 3,443 adult females who participated in the National Health and Nutrition Examination Survey from 2007 to 2012 were included. Further classification of the UI group was made into urge urinary incontinence (UUI), stress urinary incontinence (SUI), and mixed urinary incontinence (MUI). Thyroid homeostasis parameters, such as thyroid feedback quantile index (TFQI), were calculated from thyroid hormones to reflect thyroid hormone sensitivity. To examine the associations among thyroid function, thyroid homeostasis parameters and UI, we utilized weighted multivariate logistic regression and restricted cubic splines in the analysis.

**Results:** Thyroid stimulating hormone was significantly negatively correlated with UI (OR = 0.97, 95% CI: 0.96-0.99, p = 0.006), while TFQI_FT3_ showed a U-shaped association with UI (p for nonlinear = 0.014). In the analysis of subtypes, the FT3/FT4 ratio showed a negative correlation with UUI and MUI, evidenced by a J-shaped relationship, with change points identified at 0.301(p for all = 0.033, p for nonlinear =0.031; p for all = 0.010, p for nonlinear =0.005). Subgroup analysis showed consistent associations, and there was an interaction between BMI and FT3/FT4 ratio and UUI (p for interaction < 0.05).

**Conclusions:** The FT3/FT4 ratio may serve as a promising biomarker for assessing the risk of UUI and MUI. The associations among thyroid dysfunction, thyroid homeostasis, and UI in women suggest varying patterns across different UI subtypes.

## Introduction

Female urinary incontinence (UI), a prevalent pelvic floor dysfunction, is characterised by episodes of involuntary urine leakage and markedly affects quality of life, mental well-being and social interactions(1). As a result of factors such as aging, obesity and the rising prevalence of chronic conditions like hypertension and diabetes, UI has emerged as a major global public health concern. The research on UI mainly focused on the physical factors that cause pelvic floor muscle dysfunction, including obesity, constipation, fetal macrosomia history, pelvic surgery history and so on(2). The research on the relationship between endocrine and metabolic function and UI is limited.

Thyroid hormones play essential roles in energy metabolism, neuromuscular function, and the regulation of smooth muscle contraction(3,4). However, it remains unclear whether they contribute to UI through effects on bladder detrusor function, pelvic floor muscle tone, and neural conduction. Thyroid hormones influence multiple physiological systems, including the urinary system(5). Previous studies investigating the association between thyroid hormones and lower urinary tract symptoms have yielded inconsistent results across different thyroid hormones and study populations(6–9). Furthermore, the potential role of thyroid hormones in UI remains controversial(10). The existing studies focused on individual thyroid hormones, whereas the relationship between thyroid homeostasis and UI has not been elucidated.

Apart from the standard thyroid function markers, namely, free thyroxine (FT4), free triiodothyronine (FT3), thyroid stimulating hormone (TSH) and thyroglobulin (TG), newly proposed thyroid homeostasis parameters may effectively represent the dynamic equilibrium of the hypothalamic–pituitary–thyroid axis and the sensitivity of thyroid hormones(11,12). Notably, indices such as thyroid-stimulating hormone index (TSHI), thyrotroph thyroxine resistance index (TT4RI) and thyrotroph triiodothyronine resistance index (TT3RI) have been employed to assess the central sensitivity of thyroid hormones(13–15). Thyroid Feedback Quantile-based Index (TFQI) was more stable and reflected sensitivity to pituitary thyroxine, whereas a value of 0 signifies normal sensitivity(14,16). The FT3/FT4 ratio measures the conversion rate from FT4 to FT3, indicating the peripheral sensitivity of thyroid hormones(17). These combined indicators are closely associated with metabolic syndrome, chronic kidney disease, and chronic inflammatory conditions(18,19).

Our research utilised data from the National Health and Nutrition Examination Survey (NHANES) conducted between 2007 and 2012 to systematically assess the relationship among thyroid function, homeostasis indices and UI. This analysis aimed to enhance the development of customized thyroid management strategies for individuals with UI.

## Methods

### Study population

The NHANES protocol received approval from the Research Ethics Review Board of the NCHS (Protocol 2005-06 and Protocol 2011-17), and all participants provided written informed consent prior to the initiation of data collection. The current study employed publicly accessible de-identified NHANES data, and additional institutional review board approval was waived. This study focused on NHANES data obtained from 2007 to 2012. Figure 1 illustrates a flowchart that outlines the criteria for participant inclusion and exclusion. Among the total 30,442 participants from NHANES 2007–2012, exclusion applied to individuals who met the following criteria: (1) male (n = 15177) and younger than 20 years of age (n = 6208); (2) missing data on thyroid parameters (n = 4717) and UI (n = 423); and (3) missing covariate data (n = 474). The final analysis comprised a total of 3443 subjects.

**Figure 1.**
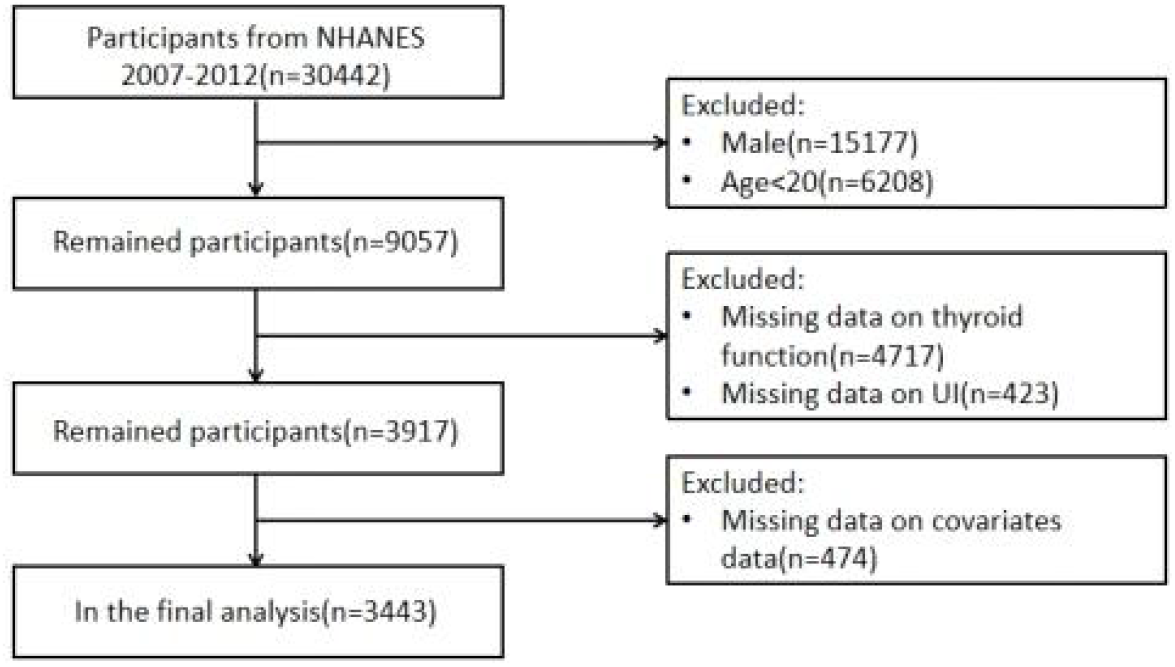
Research sample screening flow chart

### Assessment of UI

Participants responded to inquiries about UI in a Mobile Screening Center interview setting. Those who reported urine leakage during activities such as coughing or exercising within the last year were categorised as having SUI. UUI was identified in individuals experiencing involuntary urine leakage, associated with a sudden urge to urinate prior to reaching the lavatory. Individuals who met the criteria for SUI and UUI were considered MUI.

### Determination of serum thyroid function

The peripheral index of thyroid hormone sensitivity was computed as FT3/FT4 = FT3 (pg/mL)/FT4 (pmol/L), with elevated values indicating enhanced sensitivity to thyroid hormones. TSHI was calculated as ln TSH (mIU/L) + 0.1345 × FT4 (pmol/L), and the total thyroxine resistance index (TT4RI) was derived from FT4 (pmol/L) × TSH (mIU/L). Finally, the total triiodothyronine resistance index (TT3RI) was obtained from FT3 (pg/mL) × TSH (mIU/L). High values suggest a reduced sensitivity of the central nervous system to thyroid hormones. The thyroid function quality index (TFQI) was determined by utilising the population’s empirical cumulative distribution function (cdf) derived from hormone levels. TFQI_FT4_ was computed by subtracting (1 − cdf TSH) from cdf FT4. Similarly, TFQI_FT3_ was derived from cdf FT3 minus (1 − cdf TSH). Negative and positive values indicated heightened and diminished sensitivity to FT4 and FT3, respectively, while a result of 0 denotes a baseline sensitivity level.

### Assessment of covariates

Demographic information, clinical interviews, physical assessments and various laboratory data were collected in accordance with the procedures specified in the NHANES operation manuals. The sociodemographic variables comprised age (in years), race (categories included Mexican American, non-Hispanic Black, non-Hispanic White, other Hispanic and other races), educational attainment (high school or lower and beyond high school), marital status (married or cohabiting and living alone) and family poverty ratio. The health behaviours examined included smoking status (never smoked, former smoker and current smoker), alcohol intake (none, 1–5 times per month, 5–10 times per month and over 10 times per month) and exercise habits (yes or no). The health-related metrics encompassed body mass index (BMI), diabetes identified by a glycated hemoglobin level of 6.5% or above, use of diabetes medications or insulin therapy, or self-reported diabetes diagnoses. Hypertension was defined by the administration of hypotensives, a clinical diagnosis of hypertension or three consecutive readings of systolic blood pressure at or above 140 mmHg or diastolic blood pressure at or above 90 mmHg. Additionally, the dataset included information on the count of vaginal deliveries, occurrences of delivering large foetuses, history of hysterectomy and number of sexual partners.

### Statistical analysis

All analyses accounted for complex survey designs and employed appropriate survey weights. Continuous variables are expressed as mean ± standard deviation (SD), whereas categorical variables are shown as numbers (proportions). For categorical variables, Chi-square tests were utilized, and one-way ANOVA was applied to continuous variables that were normally distributed.

Weighted multivariate Logit models were used to estimate odds ratios (ORs) and 95% confidence intervals (CIs) related to the occurrence of UI and its various types. In Model 1, no adjustments were made for any variables. Model 2 included adjustments for age, education, marital status, family poverty ratio, race and BMI. In Model 3, adjustments were made for hypertension, diabetes, smoking, drinking, exercise habits, number of vaginal labour, incidence of macrosomia, history of hysterectomy and number of sexual partners.

Additionally, restricted cubic splines (RCSs) were employed to investigate the nonlinear relationship among thyroid function, thyroid homeostasis parameters and UI, considering the aforementioned covariates. Finally, subgroup analyses were conducted based on BMI, age, hypertension and hyperglycaemia. The relationships between FT3/FT4 quartiles and UI were assessed across various subgroups, with ORs, 95% CIs and p-values calculated accordingly. A p-value below 0.05 for the valid confidence interval indicated statistical significance. All statistical evaluations were conducted using R software version 4.5.1.

## Results

### Participant characteristics

A total of 3,443 individuals participated in the study and were classified into two groups based on UI status: the group with UI (n = 1,561) and the group without UI (n = 1,882). Notable differences were observed between these two groups regarding age, race, marital status, body mass index (BMI), smoking habits, alcohol intake, levels of physical activity, occurrences of hyperglycaemia and hypertension, history of hysterectomy, previous episodes of macrosomia, FT3 levels, FT3/FT4 ratio, TFQI_FT4_ values and TSHI (all p < 0.05) (Table 1). Additional analyses focusing on three types of UI revealed comparable results, with a significant difference in TG levels (Supplementary Table 1).

**Table 1.**
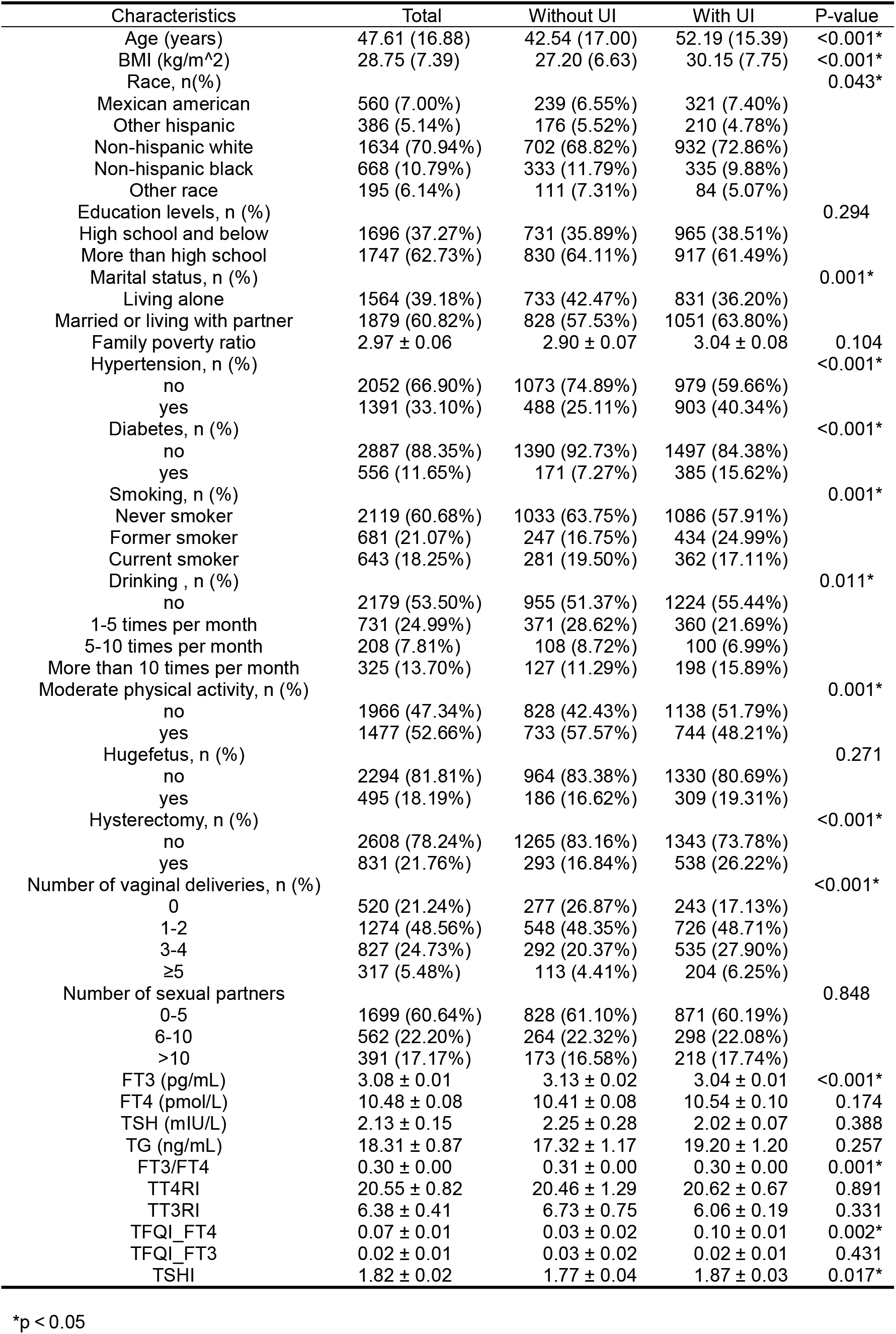
Base characteristics of participants with and without UI in the NHANES 2007–2012 cycles.

After categorizing thyroid function and thyroid homeostasis parameters into quartiles, significant differences in FT3, FT4, TSH, FT3/FT4, TT4RI, and TSHI were observed between two groups (Supplementary Table 2). Similar patterns were identified in analyses stratified by UI subtype, with further significant differences found for TG and TT3RI levels (Supplementary Table 3).

### Correlation among thyroid function, thyroid homeostasis indicators and UI

Weighted multivariate logistic regression models were utilized, as detailed in Table 2. Following adjustments for variables, it was determined that TSH exhibited an inverse relationship with UI (OR = 0.97, 95% CI = 0.96-0.99, p = 0.006). In addition, the thyroid homeostasis indices TT3RI and TT4RI were negatively correlated with UI (OR = 0.99, 95% CI = 0.98–1.00, p = 0.004; OR = 1.00, 95% CI = 0.99–1.00, p = 0.029). We found no relationship between other thyroid indicators and UI.

**Table 2.** Correlation between thyroid function/homeostasis indicators with urinary incontinence.

|  | Model 1 |  | Model 2 |  | Model 3 |  |
| --- | --- | --- | --- | --- | --- | --- |
|  | OR (95% CI) | P-value | OR (95% CI) | P-value | OR (95% CI) | P-value |
| FT3 (pg/mL) | 0.62 (0.47–0.81) | 0.001* | 0.85 (0.66–1.10) | 0.229 | 0.95 (0.80–1.12) | 0.529 |
| FT4 (pmol/L) | 1.03 (0.99–1.06) | 0.174 | 0.99 (0.95–1.03) | 0.495 | 1.01 (0.96–1.06) | 0.844 |
| TSH (mIU/L) | 0.99 (0.97–1.01) | 0.177 | 0.98 (0.96–1.00) | 0.026* | 0.97 (0.96–0.99) | 0.006* |
| TG (ng/mL) | 1.00 (1.00–1.00) | 0.357 | 1.00 (1.00–1.00) | 0.542 | 1.00 (1.00–1.01) | 0.497 |
| FT3/FT4 | 0.16 (0.05–0.55) | 0.006* | 0.80 (0.37–1.72) | 0.578 | 0.62 (0.21–1.79) | 0.384 |
| TT4RI | 1.00 (1.00–1.00) | 0.895 | 1.00 (0.99–1.00) | 0.140 | 1.00 (0.99–1.00) | 0.029* |
| TT3RI | 0.99 (0.99–1.00) | 0.111 | 0.99 (0.98–1.00) | 0.020* | 0.99 (0.98–1.00) | 0.004* |
| TFQI_FT4 | 1.60 (1.22–2.10) | 0.001* | 1.07 (0.80–1.44) | 0.640 | 1.27 (0.93–1.74) | 0.141 |
| TFQI_FT3 | 0.91 (0.72–1.15) | 0.429 | 0.93 (0.70–1.23) | 0.610 | 1.02 (0.71–1.45) | 0.931 |
| TSHI | 1.18 (1.04–1.35) | 0.016* | 1.02 (0.87–1.19) | 0.855 | 1.01 (0.85–1.20) | 0.934 |

Model 1: Non-adjusted.
Model 2: Adjusted for age, race, education level, Marital status, Family poverty ratio and BMI.
Model 3: Adjusted for age, race, education level, Marital status, Family poverty ratio, BMI, Hypertension, Diabetes, Smoking, Drinking, Moderate physical activity, Hufefetus, Hysterectomy, Number of vaginal deliveries and Number of sexual partners.
\*p < 0.05

RCS analysis revealed a nonlinear association between TFQI_FT3_ and UI (p for all = 0.032, p for nonlinear = 0.014), with the change point at 0.021 (Figure 2). No significant relationship was identified between other thyroid parameters and UI (p for all > 0.05 or p for nonlinear > 0.05).

**Figure 2.**
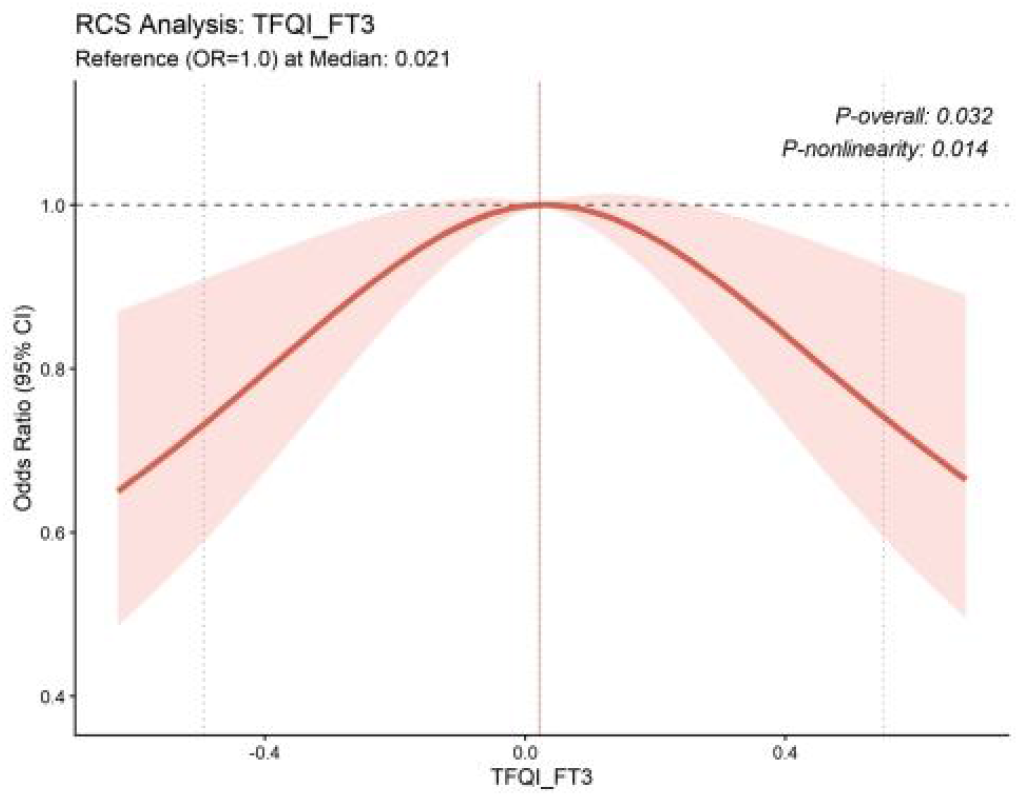
Correlation between TFQI_FT3_ and UI using a RCS Regression Model

### Correlation between thyroid function and three types of UI

Furthermore, the relationship between thyroid function and three types of UI was analyzed (Table 3, Supplementary Table 4-5). After controlling for confounding factors and grouping by quartile, FT4-Q3 was positively associated with SUI (OR = 1.43, 95% CI = 1.06-1.92, p = 0.027). Additionally, RCS analysis revealed a nonlinear U-shaped association between FT4 and SUI (P-overall = 0.045, P-nonlinearity = 0.043), with an inflection point at 10.3.And a nonlinear J-shaped association was observed between TG and UUI (P-overall = 0.013, P-nonlinearity = 0.028), with an inflection point at 11.1 (Figure 3).

**Table 3.**
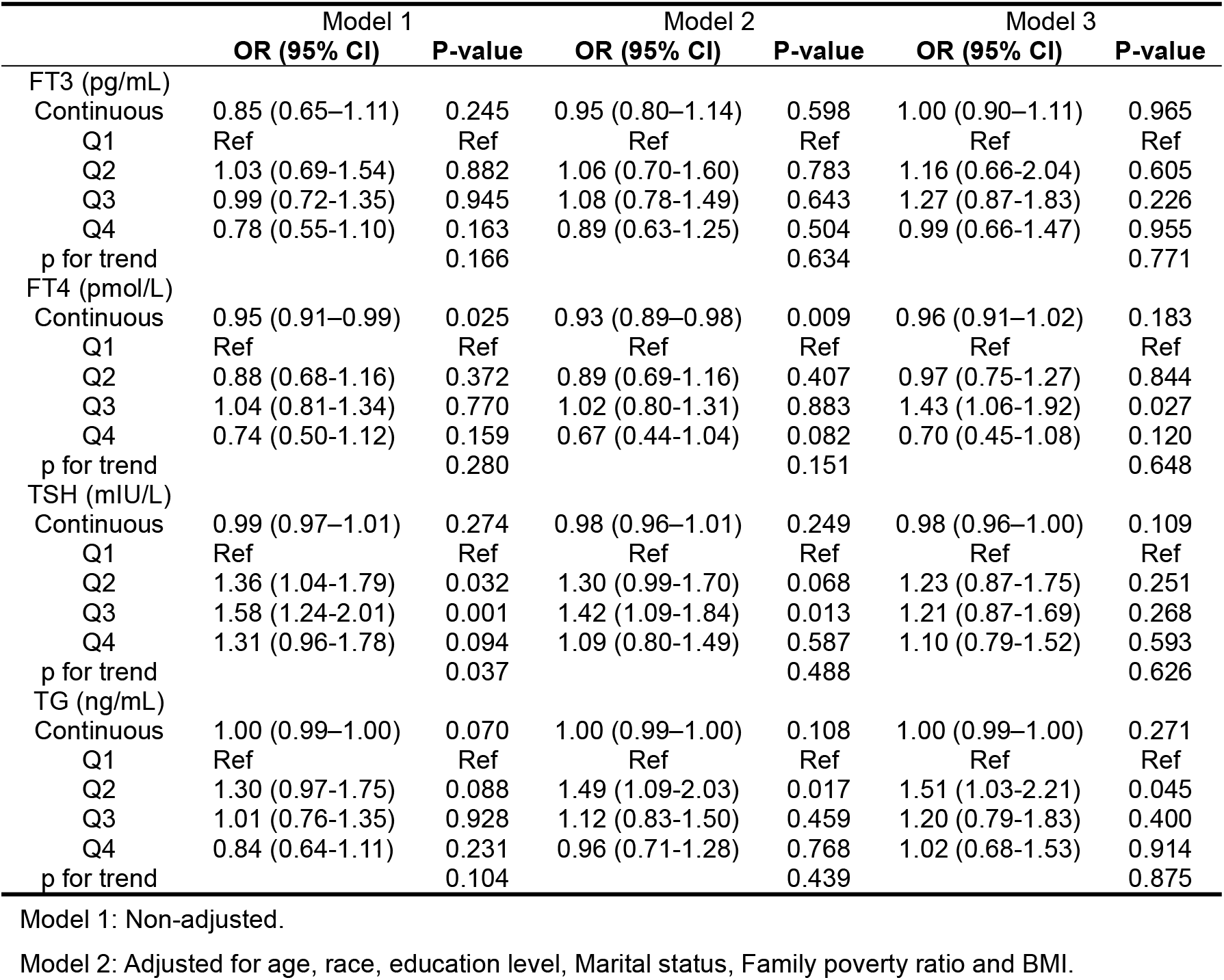
Correlation between thyroid function and stress urinary incontinence.

|  | Model 1 |  | Model 2 |  | Model 3 |  |
| --- | --- | --- | --- | --- | --- | --- |
|  | OR (95% CI) | P-value | OR (95% CI) | P-value | OR (95% CI) | P-value |
| FT3 (pg/mL) |  |  |  |  |  |  |
| Continuous | 0.85 (0.65–1.11) | 0.245 | 0.95 (0.80–1.14) | 0.598 | 1.00 (0.90–1.11) | 0.965 |
| Q1 | Ref | Ref | Ref | Ref | Ref | Ref |
| Q2 | 1.03 (0.69–1.54) | 0.882 | 1.06 (0.70–1.60) | 0.783 | 1.16 (0.66–2.04) | 0.605 |
| Q3 | 0.99 (0.72–1.35) | 0.945 | 1.08 (0.78–1.49) | 0.643 | 1.27 (0.87–1.83) | 0.226 |
| Q4 | 0.78 (0.55–1.10) | 0.163 | 0.89 (0.63–1.25) | 0.504 | 0.99 (0.66–1.47) | 0.955 |
| p for trend |  | 0.166 |  | 0.634 |  | 0.771 |
| FT4 (pmol/L) |  |  |  |  |  |  |
| Continuous | 0.95 (0.91–0.99) | 0.025 | 0.93 (0.89–0.98) | 0.009 | 0.96 (0.91–1.02) | 0.183 |
| Q1 | Ref | Ref | Ref | Ref | Ref | Ref |
| Q2 | 0.88 (0.68–1.16) | 0.372 | 0.89 (0.69–1.16) | 0.407 | 0.97 (0.75–1.27) | 0.844 |
| Q3 | 1.04 (0.81–1.34) | 0.770 | 1.02 (0.80–1.31) | 0.883 | 1.43 (1.06–1.92) | 0.027 |
| Q4 | 0.74 (0.50–1.12) | 0.159 | 0.67 (0.44–1.04) | 0.082 | 0.70 (0.45–1.08) | 0.120 |
| p for trend |  | 0.280 |  | 0.151 |  | 0.648 |
| TSH (mIU/L) |  |  |  |  |  |  |
| Continuous | 0.99 (0.97–1.01) | 0.274 | 0.98 (0.96–1.01) | 0.249 | 0.98 (0.96–1.00) | 0.109 |
| Q1 | Ref | Ref | Ref | Ref | Ref | Ref |
| Q2 | 1.36 (1.04–1.79) | 0.032 | 1.30 (0.99–1.70) | 0.068 | 1.23 (0.87–1.75) | 0.251 |
| Q3 | 1.58 (1.24–2.01) | 0.001 | 1.42 (1.09–1.84) | 0.013 | 1.21 (0.87–1.69) | 0.268 |
| Q4 | 1.31 (0.96–1.78) | 0.094 | 1.09 (0.80–1.49) | 0.587 | 1.10 (0.79–1.52) | 0.593 |
| p for trend |  | 0.037 |  | 0.488 |  | 0.626 |
| TG (ng/mL) |  |  |  |  |  |  |
| Continuous | 1.00 (0.99–1.00) | 0.070 | 1.00 (0.99–1.00) | 0.108 | 1.00 (0.99–1.00) | 0.271 |
| Q1 | Ref | Ref | Ref | Ref | Ref | Ref |
| Q2 | 1.30 (0.97–1.75) | 0.088 | 1.49 (1.09–2.03) | 0.017 | 1.51 (1.03–2.21) | 0.045 |
| Q3 | 1.01 (0.76–1.35) | 0.928 | 1.12 (0.83–1.50) | 0.459 | 1.20 (0.79–1.83) | 0.400 |
| Q4 | 0.84 (0.64–1.11) | 0.231 | 0.96 (0.71–1.28) | 0.768 | 1.02 (0.68–1.53) | 0.914 |
| p for trend |  | 0.104 |  | 0.439 |  | 0.875 |

Model 1: Non-adjusted.
Model 2: Adjusted for age, race, education level, Marital status, Family poverty ratio and BMI.

**Figure 3.**
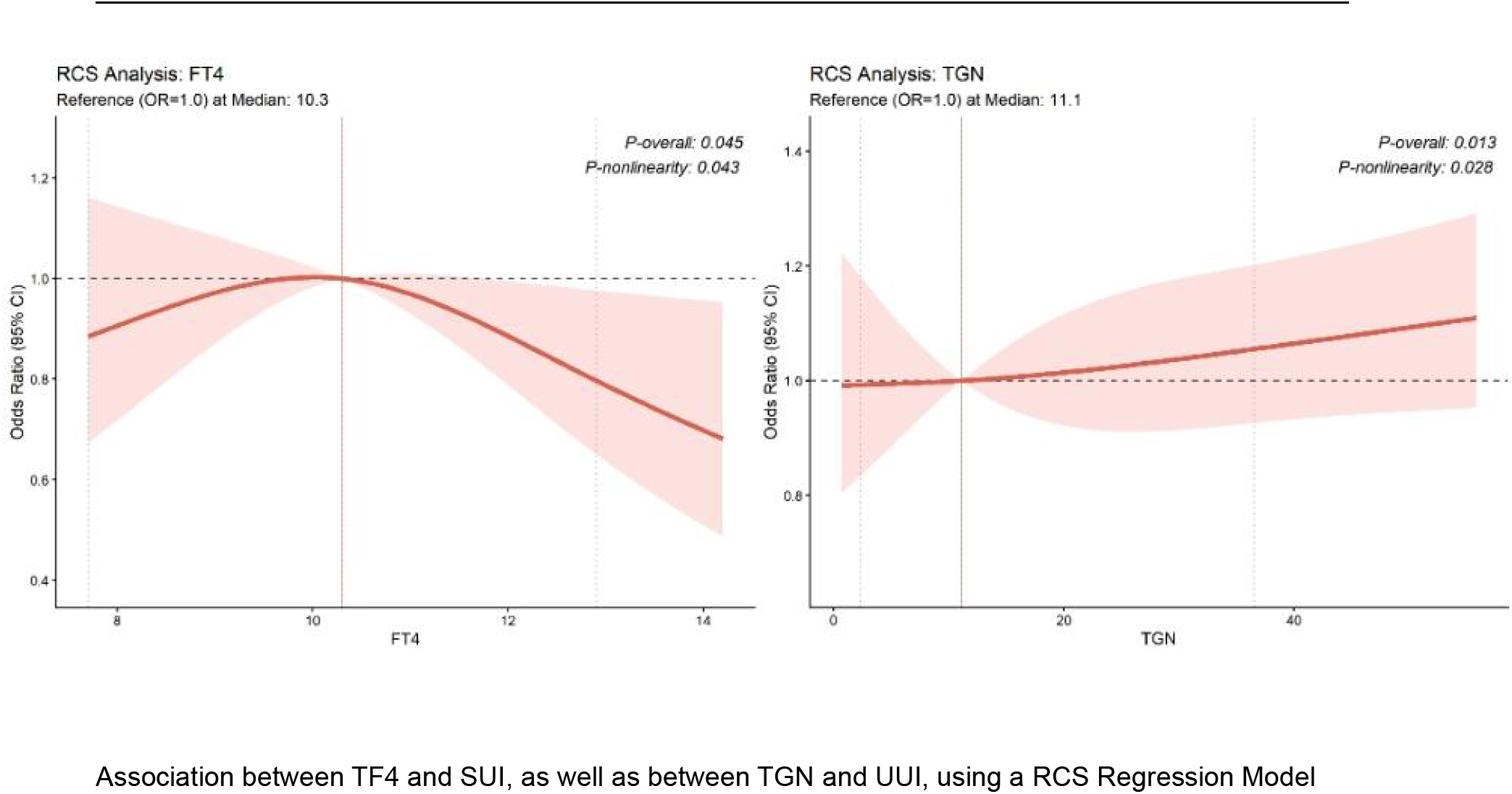
Association between TF4 and SUI, as well as between TGN and UUI, using a RCS Regression Model

### Association between thyroid homeostasis indicators and three types of UI

After controlling for covariates, the analysis revealed a negative correlation between the FT3/FT4 ratio of thyroid homeostasis and UUI and MUI when stratified into quartiles (Table 4, Supplementary Table 6). For UUI, the OR and 95% CI for the FT3/FT4 categories from lowest to highest were as follows: 1.00 (reference), OR = 0.63; 95% CI = 0.43–0.92, p = 0.025, OR = 0.64; 95% CI = 0.46–0.88, p = 0.011, and OR = 0.65; and 95% CI = 0.44–0.98, p = 0.049, with a p trend = 0.045. For MUI, the OR and 95% CI across the FT3/FT4 categories from lowest to highest were 1.00 (reference), OR = 0.63; 95% CI = 0.44–0.91, p = 0.021, OR = 0.59; 95% CI = 0.42–0.83, p = 0.007, and OR = 0.64; and 95% CI = 0.44–0.92, p = 0.025, with a p trend = 0.009.

**Table 4.**
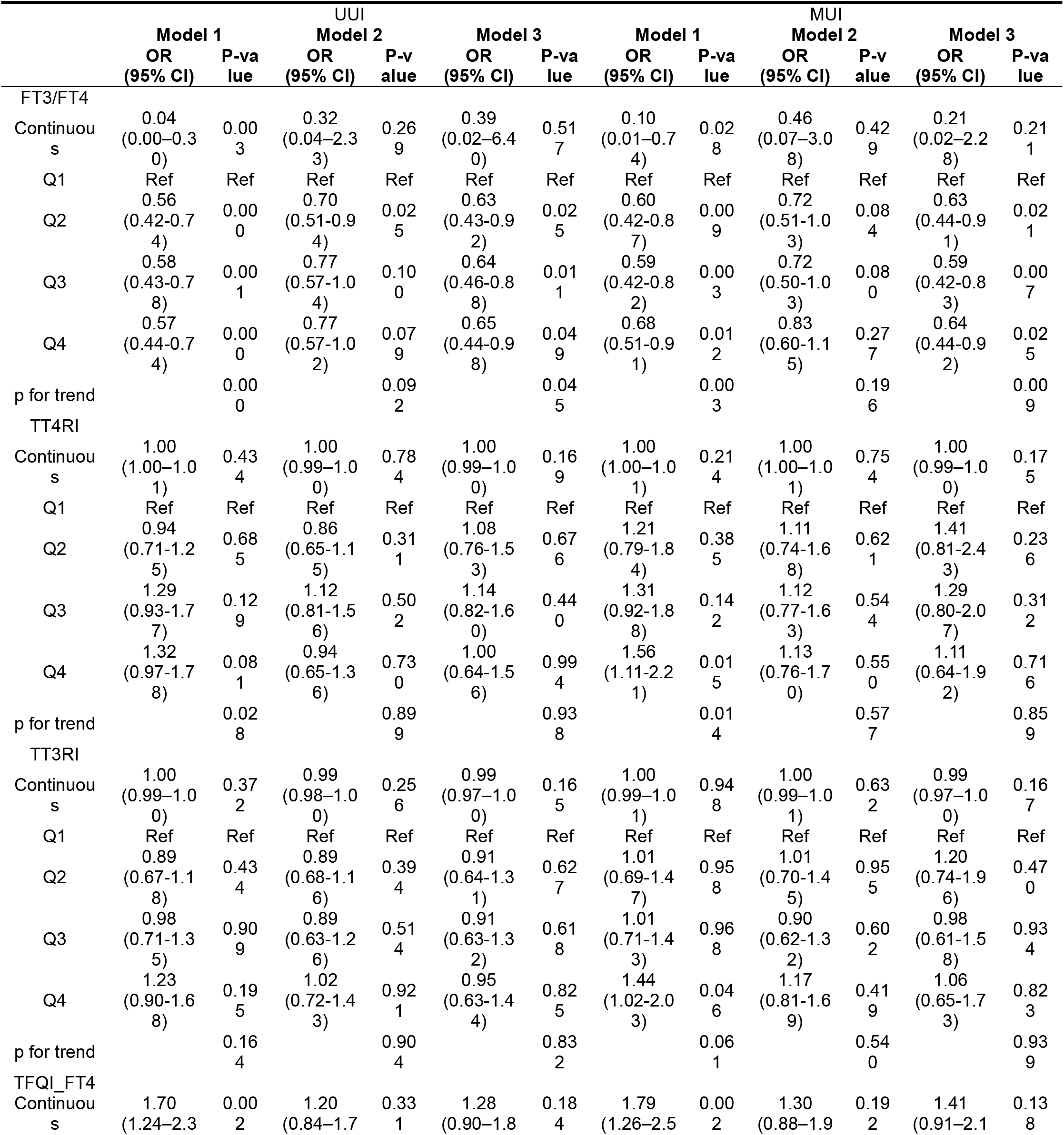
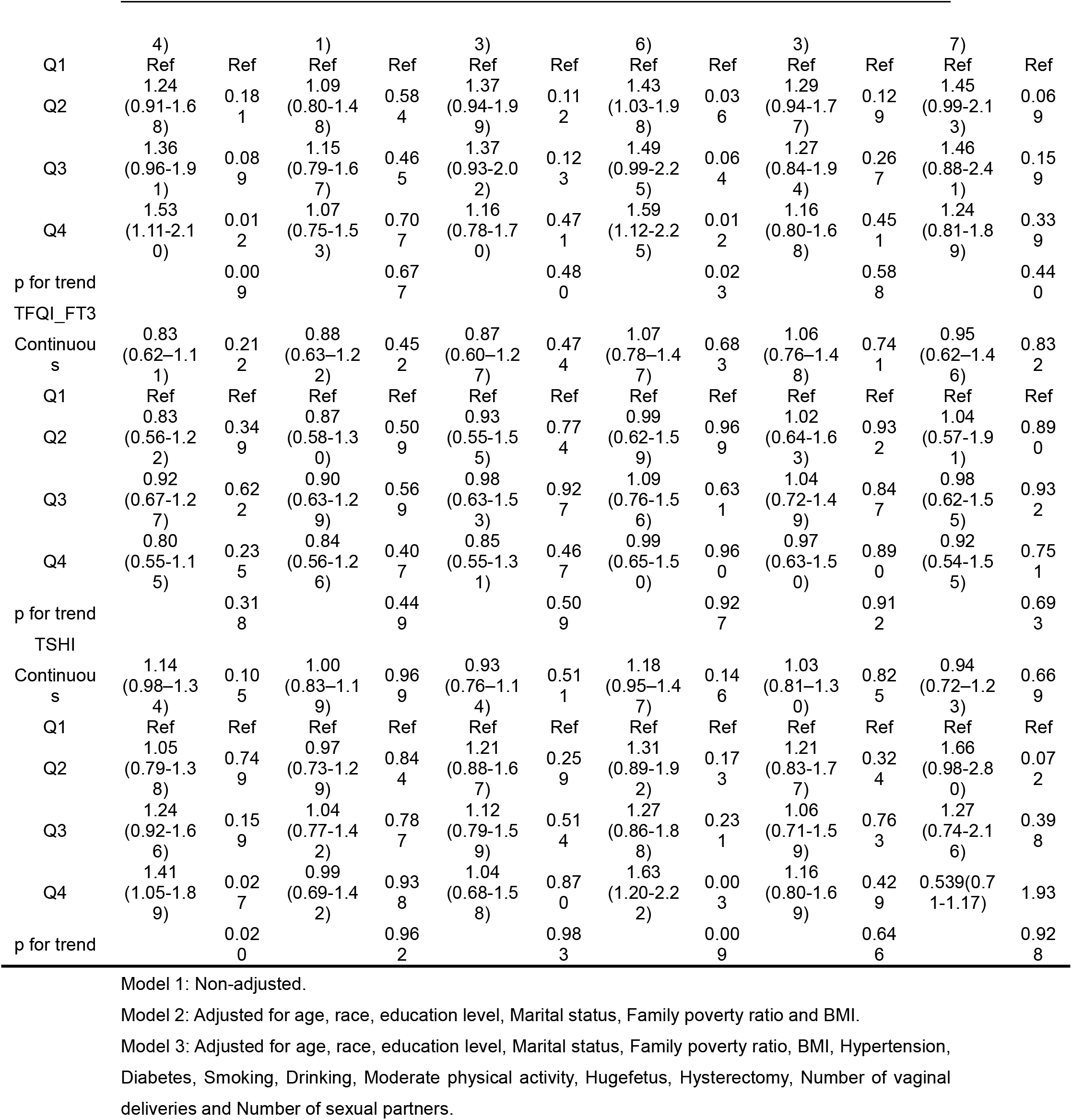
Association between thyroid homeostasis indicators with UUI and MUI

In addition, there was a J-shaped nonlinear relationship between FT3/FT4 ratio and UUI and MUI (p for all = 0.033, p for nonlinear =0.031; p for all = 0.010, p for nonlinear =0.005), with the same inflection point at 0.301(Figure 4).

**Figure 4.**
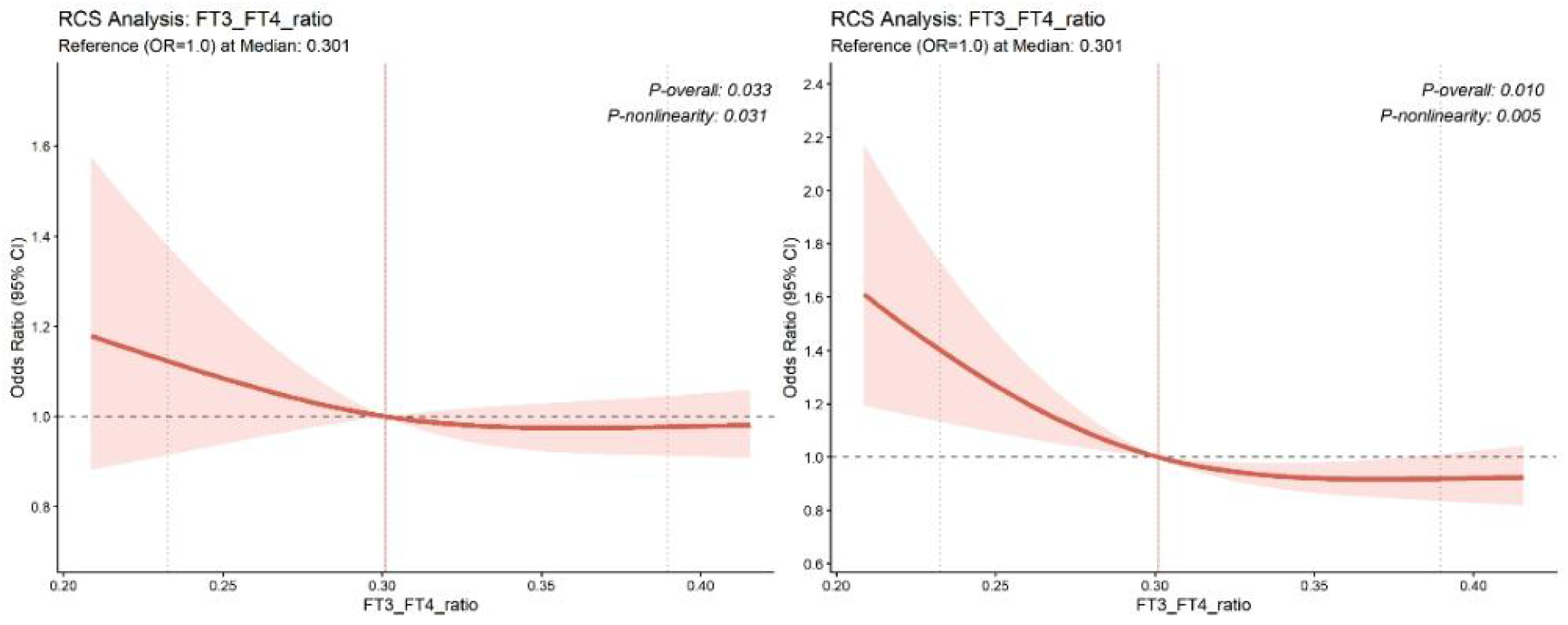
Association between FT3/FT4 ratio with UUI and MUI using a RCS Regression Model

### subgroup analysis

To further study the correlation between the FT3/FT4 ratio and UUI and MUI, we performed a subgroup analysis stratified by BMI, age, hypertension and hyperglycaemia (Figure 5). These findings suggest that the inverse associations between the FT3/FT4 ratio and the risks of UUI and MUI remained consistent across most subgroups. A significant interaction was observed between the FT3/FT4 ratio and BMI for UUI risk (P for interaction < 0.05), whereas no significant interactions were detected for age, hypertension, or hyperglycemia (all P for interaction > 0.05).

**Figure 5.**
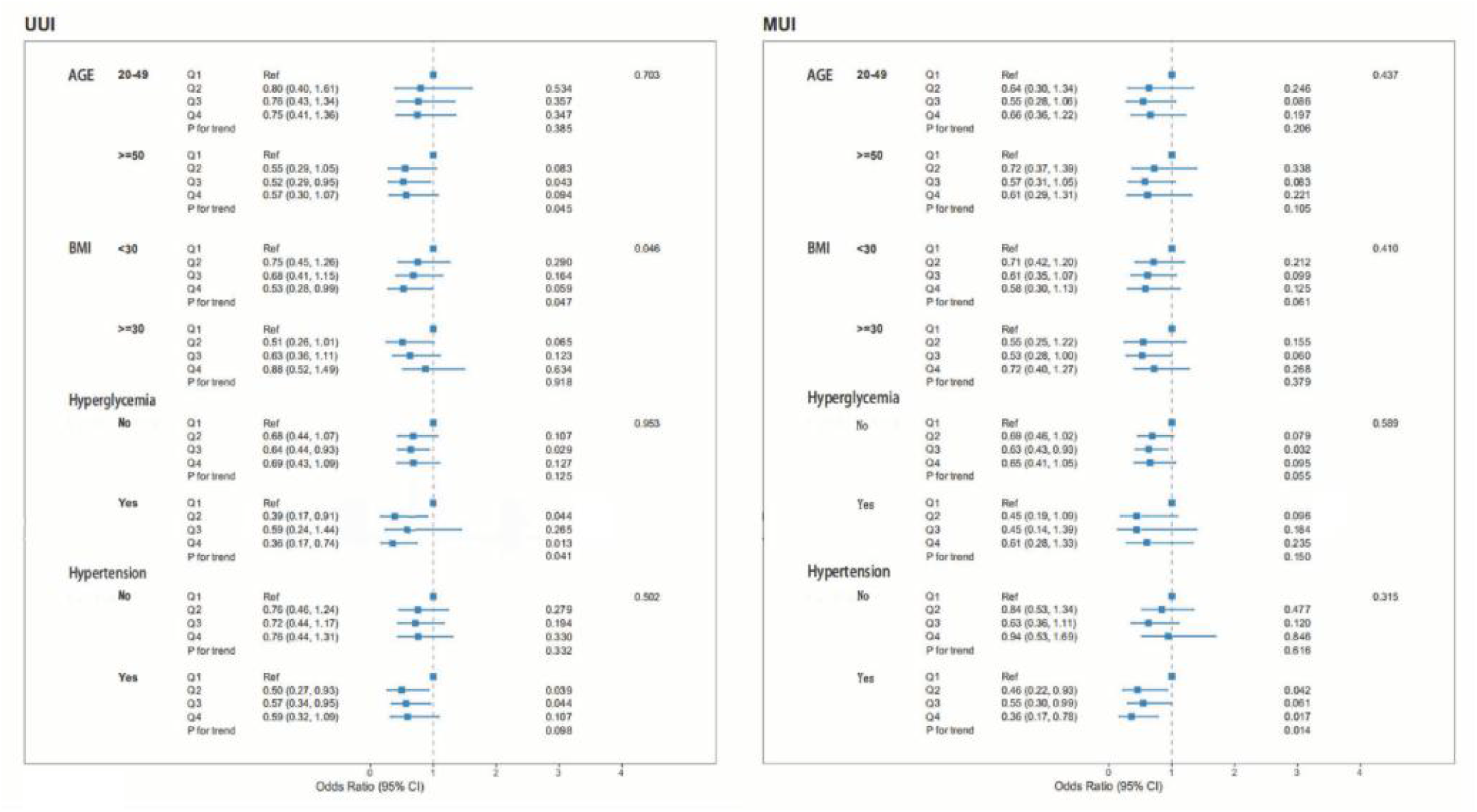
Subgroup analyses between FT3/FT4 ratio and UUI, MUI stratified by age, BMI, hyperglycemia and hypertension.

## Discussion

To our knowledge, this is the first study to investigate the associations between thyroid homeostasis parameters and female UI. Our findings suggest that thyroid function and thyroid homeostasis parameters exhibit distinct associations across different types of UI. Significant associations were observed between TSH and overall UI. In addition, specific associations were identified between the FT3/FT4 ratio and both UUI and MUI. RCS analyses suggested that the effects of thyroid dysfunction on lower urinary tract function may be more complex than a simple linear relationship. These findings provide new insights into the potential role of thyroid homeostasis in the pathogenesis of female UI.

Firstly, in our study, age, race, marital status, BMI, smoking, drinking, physical activity, hyperglycaemia, hypertension, hysterectomy and history of macrosomia were statistically different between the UI and without UI groups, which was consistent with previous studies(20,21). The study found an inverse association between TSH and UI, which suggested that a mild tendency to hyperthyroidism, even within the clinically normal range, may affect female pelvic floor function. This is consistent with the previous epidemiological study(22). In addition, a case report indicated that a female with SUI and hyperthyroidism experienced alleviation of lower urinary tract symptoms after thyroid function treatment(23). A variety of mechanisms may be involved. Firstly, thyroid hormones can regulate sympathetic nerve activity, detrusor contractility and bladder sensory function(24–27). Abnormal thyroid function may lead to overactive bladder and decreased urethral sphincter stability, increasing the risk of UI. Secondly, heightened thirst and water intake increase kidney and bladder burden(28). Thyroid hormones are involved in skeletal muscle metabolism and collagen tissue homeostasis(29), and long-term thyroid dysfunction may compromise the support structures of the pelvic floor and further promote the occurrence of UI.

Compared with peripheral thyroid hormones, TSH and thyroid homeostasis parameters are better indicators of subtle dysregulation of the thyroid axis. TT3RI and TT4RI are regarded as significant markers of the homeostatic regulation of the thyroid axis, with elevated values suggesting reduced sensitivity of the central nervous system to feedback from thyroid hormones(13–15). Our research revealed a significant negative correlation among TT3RI, TT4RI and UI, consistent with previous findings that TSH exhibited a negative correlation with UI. Furthermore, even when FT3 and FT4 levels remain within the normal range, alterations in the sensitivity of thyroid axis feedback may indicate mild abnormalities in thyroid homeostasis. Therefore, TT3RI and TT4RI may be more sensitive than traditional thyroid hormone indicators in reflecting the endocrine changes associated with UI.

Notably, the RCS analysis conducted in this study revealed a U-shaped relationship between TFQI_FT3_ and UI, indicating that insufficient or excessive thyroid feedback sensitivity may increase the risk of UI. This nonlinear relationship suggested that maintaining an appropriate thyroid axis feedback balance may be critical for the stability of pelvic floor function. The finding was biologically plausible and consistent with the physiological functions of thyroid hormones. Excessive thyroid hormone levels may increase detrusor nerve excitability and promote bladder overactivity, whereas insufficient hormone levels may impair smooth muscle function and neural conduction. Consequently, both hyperthyroid and hypothyroid states may contribute to lower urinary tract dysfunction, providing a potential explanation for the observed nonlinear associations and the heterogeneous findings reported in previous studies(7,8,30). The findings further support the important role of ’thyroid homeostasis imbalance’ rather than ’increased or decreased thyroid hormone’ in UI.

Our analysis of different types of UI revealed a U-shaped relationship between FT4 and SUI, which was not present in other types of UI. This may be because the core mechanism of SUI is related to the damage of the pelvic floor support structure and the reduction in urethral closure pressure(31). Compared with other types of UI, SUI is more dependent on the pelvic floor’s support structure and muscle function. Abnormal FT4 levels may affect skeletal muscle metabolism, collagen synthesis and oestrogen-related pathways, thereby altering the elasticity and muscle function of pelvic floor tissue and increasing the risk of SUI(32,33). Additionally, a J-shaped relationship between TG and UUI was observed in this study. TG serves as a storage carrier and synthetic precursor of thyroid hormone, and its elevation may reflect underlying thyroid inflammation(34,35). Inflammation may influence bladder sensory sensitivity via autonomic nerve dysfunction and chronic low-grade inflammation, facilitating the occurrence of UUI. These results suggest that different types of UI may operate through distinct endocrine mechanisms.

A key discovery of this research was the consistent inverse correlation observed between the FT3/FT4 ratio and UUI and MUI, which showed a clear J-shaped association. Typically, the FT3/FT4 ratio is viewed as a marker for peripheral deiodinase activity and the effectiveness of T4 conversion to T3. A low FT3/FT4 ratio indicates a decrease in the peripheral conversion capacity of thyroid hormones, which may reflect the underutilisation of thyroid hormones at the tissue level(17,36). The correlation between the ratio and various endocrine and metabolic diseases has been established(37–39), resulting in its identification as a potential biomarker for muscle atrophy(40). Research indicates that a low FT3/FT4 ratio can serve as an alternative marker for muscle atrophy in individuals suffering from advanced non-small cell lung cancer(41). In older adults with heart failure, a low FT3/FT4 ratio is associated with frailty, malnutrition, inflammation and heightened mortality risk(42). Therefore, a reduction in the FT3/FT4 ratio may adversely affect the function of pelvic floor muscles and bladder nerves, leading to an elevated risk of UUI and MUI. RCS analysis revealed a J-shaped association, indicating that a low FT3/FT4 ratio may represent a critical threshold for a significant risk increase, with the change point noted at 0.301. This finding provides valuable insights for the clinical assessment of the relationship between thyroid hormone turnover efficiency and pelvic floor functionality.

Furthermore, subgroup analysis revealed an interaction effect of BMI on the relationship between the FT3/FT4 ratio and UUI, but age, hypertension and hyperglycaemia had no significant interaction effect. Obesity may aggravate pelvic floor dysfunction by increasing abdominal pressure, promoting a chronic inflammatory state and affecting thyroid hormone metabolism(43). Therefore, abnormal thyroid homeostasis may have a more significant effect on UUI in people with high BMI than in those with low BMI. This suggests that increased clinical attention should be directed towards the synergistic effect between thyroid function and UI in obese females.

A notable strength of our study is that it was based on NHANES, a nationally representative database, and used a complex sampling weighted analysis, which increases the representativeness and reliability of the results. This study not only analysed traditional thyroid function indicators but also systematically included thyroid homeostasis markers such as TFQI, TT3RI, TT4RI and FT3/FT4 for a comprehensive assessment. Nonetheless, our study had limitations that warrant consideration. Firstly, the cross-sectional nature of this study prevented us from establishing a causal link between thyroid dysfunction and UI. Secondly, despite the adjustment for multiple confounders, their potential effects remain significant, and future investigations should include a wide range of adjustment factors.

## Conclusion

In conclusion, this research represents the first population-based investigation into the relationship among thyroid function, homeostasis parameters and the prevalence of UI and its subtypes. An imbalance in thyroid homeostasis may contribute to the occurrence of UI, particularly the FT3/FT4 ratio, which may be a potential biomarker for the risk of UUI and MUI. Prospective and mechanistic studies are necessary to clarify the causal role and potential biological mechanisms.

## Supplementary Information

Supplementary Table 1. Basic characteristics of participants with different types of UI.

Supplementary Table 2. The quartile distribution of thyroid function/homeostasis parameters in participants with and without UI.

Supplementary Table 3. The quartile distribution of thyroid function/homeostasis parameters in participants with different types of UI.

Supplementary Table 4. Correlation between thyroid function and UUI.

Supplementary Table 5. Correlation between thyroid function and MUI.

Supplementary Table 6. Association between thyroid homeostasis parameters with SUI.

## Author Contributions

Kaixian Deng conceived the presented idea. Junchao Zhang performed the computations and manuscript writing. The acquisition of data involved Qian Yang, Jinfa Huang, Huan Yang, Ruolan Li and Kirtchhoof Tevorn. All authors participated in the article and endorsed the submitted version.

## Data availability

The information utilized in this research originates from a publicly available database found at https://www.cdc.gov/nchs/nhanes/index.htm, which is accessible to all through the links included in the document.

## Declarations

The research was conducted in accordance with the Declaration of Helsinki and obtained consent from the NCHS Ethics Review Board. All subjects gave their written informed consent to participate.

## Acknowledgement

The authors would like to thank all individuals who contributed to this study.

