## Supplemental Table1-6 for "Association between thyroid function and thyroid homeostasis parameters and female urinary incontinence: A population-based study": Supplementary Table 1.pdf

Supplementary Table 1. Basic characteristics of participants with different types of urinary incontinence in the NHANES 2007–2012 cycles.

| Characteristics | Total | SUI | UUI | MUI | Without UI | P-value |
|---|---|---|---|---|---|---|
| Age (years) | 47.61 ± 0.56 | 50.02 ± 0.81 | 55.55 ± 1.39 | 53.50 ± 0.70 | 42.54 ± 0.70 | <0.001* |
| BMI (kg/m^2) | 28.75 ± 0.18 | 29.59 ± 0.38 | 30.37 ± 0.55 | 30.92 ± 0.50 | 27.20 ± 0.25 | <0.001* |
| Race, n(%) | | | | | | <0.001* |
| Mexican american | 560 (7.00%) | 171 (8.64%) | 48 (5.07%) | 102 (6.95%) | 239 (6.55%) | |
| Other hispanic | 386 (5.14%) | 91 (5.02%) | 41 (4.18%) | 78 (4.80%) | 176 (5.52%) | |
| Non-hispanic white | 1634 (70.94%) | 449 (72.99%) | 192 (70.04%) | 291 (74.54%) | 702 (68.82%) | |
| Non-hispanic black | 668 (10.79%) | 112 (6.89%) | 130 (18.38%) | 93 (9.06%) | 333 (11.79%) | |
| Other race | 195 (6.14%) | 49 (6.46%) | 11 (2.33%) | 24 (4.65%) | 111 (7.31%) | |
| Education levels, n (%) | | | | | | 0.286 |
| High school and below | 1696 (37.27%) | 419 (36.44%) | 212 (38.22%) | 334 (42.09%) | 731 (35.89%) | |
| More than high school | 1747 (62.73%) | 453 (63.56%) | 210 (61.78%) | 254 (57.91%) | 830 (64.11%) | |
| Marital status, n (%) | | | | | | 0.001* |
| Living alone | 1564 (39.18%) | 327 (30.89%) | 224 (44.39%) | 280 (39.38%) | 733 (42.47%) | |
| Married or living with partner | 1879 (60.82%) | 545 (69.11%) | 198 (55.61%) | 308 (60.62%) | 828 (57.53%) | |
| Family poverty ratio | 2.97 ± 0.06 | 3.21 ± 0.10 | 3.01 ± 0.12 | 2.78 ± 0.14 | 2.90 ± 0.07 | 0.063 |
| Hypertension, n (%) | | | | | | <0.001* |
| no | 2052 (66.90%) | 517 (65.02%) | 184 (50.72%) | 278 (56.92%) | 1073 (74.89%) | |
| yes | 1391 (33.10%) | 355 (34.98%) | 238 (49.28%) | 310 (43.08%) | 488 (25.11%) | |
| Diabetes, n (%) | | | | | | <0.001* |
| no | 2887 (88.35%) | 737 (88.13%) | 328 (84.73%) | 432 (78.06%) | 1390 (92.73%) | |
| yes | 556 (11.65%) | 135 (11.87%) | 94 (15.27%) | 156 (21.94%) | 171 (7.27%) | |
| Smoking, n (%) | | | | | | 0.004* |
| Never smoker | 2119 (60.68%) | 518 (62.21%) | 248 (57.50%) | 320 (51.19%) | 1033 (63.75%) | |
| Former smoker | 681 (21.07%) | 195 (22.72%) | 101 (24.26%) | 138 (29.15%) | 247 (16.75%) | |
| Current smoker | 643 (18.25%) | 159 (15.07%) | 73 (18.23%) | 130 (19.67%) | 281 (19.50%) | |
| Drinking , n (%) | | | | | | 0.005* |
| no | 2179 (53.50%) | 524 (49.91%) | 279 (59.84%) | 421 (61.48%) | 955 (51.37%) | |
| 1-5 times per month | 731 (24.99%) | 181 (23.88%) | 82 (20.56%) | 97 (18.88%) | 371 (28.62%) | |
| 5-10 times per month | 208 (7.81%) | 53 (7.49%) | 18 (4.71%) | 29 (7.68%) | 108 (8.72%) | |
| More than 10 times per month | 325 (13.70%) | 114 (18.72%) | 43 (14.88%) | 41 (11.95%) | 127 (11.29%) | |
| Moderate physical activity, n (%) | | | | | | 0.003* |

| Characteristics | Total | SUI | UUI | MUI | Without UI | P-value |
|---|---|---|---|---|---|---|
| no | 1966 (47.34%) | 492 (47.84%) | 270 (58.38%) | 376 (53.83%) | 828 (42.43%) | |
| yes | 1477 (52.66%) | 380 (52.16%) | 152 (41.62%) | 212 (46.17%) | 733 (57.57%) | |
| Hugefetus, n (%) | | | | | | 0.073 |
| no | 2294 (81.81%) | 620 (83.81%) | 283 (74.07%) | 427 (79.67%) | 964 (83.38%) | |
| yes | 495 (18.19%) | 136 (16.19%) | 73 (25.93%) | 100 (20.33%) | 186 (16.62%) | |
| Hysterectomy, n (%) | | | | | | <0.001* |
| no | 2608 (78.24%) | 680 (78.49%) | 285 (71.96%) | 378 (67.34%) | 1265 (83.16%) | |
| yes | 831 (21.76%) | 192 (21.51%) | 137 (28.04%) | 209 (32.66%) | 293 (16.84%) | |
| Number of vaginal deliveries, n (%) | | | | | | <0.001* |
| 0 | 520 (21.24%) | 120 (16.68%) | 63 (25.03%) | 60 (12.89%) | 277 (26.87%) | |
| 1-2 | 1274 (48.56%) | 352 (50.74%) | 146 (40.10%) | 228 (50.88%) | 548 (48.35%) | |
| 3-4 | 827 (24.73%) | 233 (27.56%) | 118 (27.12%) | 184 (28.94%) | 292 (20.37%) | |
| ≥5 | 317 (5.48%) | 82 (5.01%) | 50 (7.75%) | 72 (7.29%) | 113 (4.41%) | |
| Number of sexual partners | | | | | | 0.945 |
| 0-5 | 1699 (60.64%) | 440 (59.76%) | 159 (59.24%) | 272 (61.42%) | 828 (61.10%) | |
| 6-10 | 562 (22.20%) | 144 (21.28%) | 57 (23.58%) | 97 (22.58%) | 264 (22.32%) | |
| >10 | 391 (17.17%) | 114 (18.96%) | 42 (17.18%) | 62 (16.00%) | 173 (16.58%) | |
| FT3 (pg/mL) | 3.08 ± 0.01 | 3.06 ± 0.02 | 3.00 ± 0.02 | 3.05 ± 0.02 | 3.13 ± 0.02 | <0.001* |
| FT4 (pmol/L) | 10.48 ± 0.08 | 10.30 ± 0.10 | 10.84 ± 0.25 | 10.73 ± 0.16 | 10.41 ± 0.08 | 0.011* |
| TSH (mIU/L) | 2.13 ± 0.15 | 2.01 ± 0.09 | 1.89 ± 0.10 | 2.13 ± 0.14 | 2.25 ± 0.28 | 0.384 |
| TG (ng/mL) | 18.31 ± 0.87 | 15.56 ± 0.96 | 25.09 ± 4.50 | 21.21 ± 2.04 | 17.32 ± 1.17 | 0.012* |
| FT3/FT4 | 0.30 ± 0.00 | 0.31 ± 0.00 | 0.29 ± 0.01 | 0.30 ± 0.00 | 0.31 ± 0.00 | 0.002* |
| TT4RI | 20.55 ± 0.82 | 19.90 ± 0.73 | 20.05 ± 1.00 | 22.19 ± 1.63 | 20.46 ± 1.29 | 0.636 |
| TT3RI | 6.38 ± 0.41 | 6.06 ± 0.27 | 5.63 ± 0.29 | 6.35 ± 0.38 | 6.73 ± 0.75 | 0.331 |
| TFQI_FT4 | 0.07 ± 0.01 | 0.07 ± 0.02 | 0.10 ± 0.03 | 0.13 ± 0.02 | 0.03 ± 0.02 | 0.010* |
| TFQI_FT3 | 0.02 ± 0.01 | 0.03 ± 0.02 | -0.04 ± 0.03 | 0.03 ± 0.02 | 0.03 ± 0.02 | 0.090 |
| TSHI | 1.82 ± 0.02 | 1.86 ± 0.03 | 1.84 ± 0.05 | 1.90 ± 0.06 | 1.77 ± 0.04 | 0.108* |
