## Supplemental Table1-6 for "Association between thyroid function and thyroid homeostasis parameters and female urinary incontinence: A population-based study": Supplementary Table 2.pdf

Supplementary Table 2. The quartile distribution of thyroid function/homeostasis parameters in participants with and without UI in the NHANES 2007–2012 cycles.

| Characteristics | Total | Without UI | With UI | P-value |
|---|---|---|---|---|
| FT3 (pg/mL) | | | | |
| Q1 | 902 (24.11%) | 374 (20.87%) | 528 (27.06%) | 0.001* |
| Q2 | 825 (24.83%) | 360 (23.49%) | 465 (26.04%) | |
| Q3 | 1020 (30.62%) | 484 (31.81%) | 536 (29.54%) | |
| Q4 | 696 (20.44%) | 343 (23.83%) | 353 (17.36%) | |
| FT4 (pmol/L) | | | | |
| Q1 | 1223 (29.49%) | 559 (29.68%) | 664 (29.31%) | 0.045* |
| Q2 | 1040 (30.70%) | 479 (32.87%) | 561 (28.72%) | |
| Q3 | 660 (21.58%) | 303 (21.75%) | 357 (21.43%) | |
| Q4 | 520 (18.23%) | 220 (15.69%) | 300 (20.53%) | |
| TSH (mIU/L) | | | | |
| Q1 | 866 (23.94%) | 426 (27.45%) | 440 (20.77%) | 0.010* |
| Q2 | 856 (24.56%) | 408 (25.01%) | 448 (24.15%) | |
| Q3 | 861 (27.82%) | 381 (25.85%) | 480 (29.61%) | |
| Q4 | 860 (23.68%) | 346 (21.70%) | 514 (25.47%) | |
| TG (ng/mL) | | | | |
| Q1 | 862 (25.89%) | 384 (25.64%) | 478 (26.12%) | 0.650 |
| Q2 | 860 (24.81%) | 411 (24.98%) | 449 (24.65%) | |
| Q3 | 860 (24.89%) | 383 (26.09%) | 477 (23.80%) | |
| Q4 | 861 (24.41%) | 383 (23.29%) | 478 (25.43%) | |
| FT3/FT4 | | | | |
| Q1 | 901 (27.71%) | 376 (23.92%) | 525 (31.15%) | 0.011* |
| Q2 | 850 (24.94%) | 394 (25.96%) | 456 (24.01%) | |
| Q3 | 874 (24.83%) | 395 (26.02%) | 479 (23.76%) | |
| Q4 | 818 (22.52%) | 396 (24.10%) | 422 (21.09%) | |
| TT4RI | | | | |
| Q1 | 861 (23.11%) | 419 (25.85%) | 442 (20.63%) | 0.020* |
| Q2 | 861 (25.18%) | 409 (26.50%) | 452 (23.98%) | |
| Q3 | 860 (26.51%) | 386 (24.91%) | 474 (27.97%) | |
| Q4 | 861 (25.20%) | 347 (22.74%) | 514 (27.42%) | |
| TT3RI | | | | |
| Q1 | 861 (23.74%) | 415 (26.18%) | 446 (21.54%) | 0.059 |
| Q2 | 862 (24.76%) | 410 (25.99%) | 452 (23.65%) | |
| Q3 | 859 (26.76%) | 384 (24.52%) | 475 (28.79%) | |
| Q4 | 861 (24.73%) | 352 (23.31%) | 509 (26.02%) | |
| TFQI_FT4 | | | | |
| Q1 | 861 (24.81%) | 415 (27.09%) | 446 (22.76%) | 0.062 |
| Q2 | 861 (25.17%) | 400 (26.41%) | 461 (24.04%) | |
| Q3 | 861 (24.44%) | 386 (23.62%) | 475 (25.18%) | |
| Q4 | 860 (25.58%) | 360 (22.88%) | 500 (28.02%) | |
| TFQI_FT3 | | | | |
| Q1 | 863 (26.38%) | 391 (25.60%) | 472 (27.09%) | 0.851 |
| Q2 | 860 (23.89%) | 406 (24.29%) | 454 (23.53%) | |
| Q3 | 859 (23.60%) | 383 (24.26%) | 476 (23.00%) | |

| Characteristics | Total | Without UI | With UI | P-value |
|---|---|---|---|---|
| Q4 | 861 (26.13%) | 381 (25.86%) | 480 (26.38%) | |
| TSHI | | | | |
| Q1 | 861 (22.93%) | 425 (25.83%) | 436 (20.30%) | 0.006* |
| Q2 | 863 (25.75%) | 410 (27.05%) | 453 (24.58%) | |
| Q3 | 858 (26.41%) | 376 (24.37%) | 482 (28.26%) | |
| Q4 | 861 (24.91%) | 350 (22.74%) | 511 (26.87%) | |
