## Supplemental Table1-6 for "Association between thyroid function and thyroid homeostasis parameters and female urinary incontinence: A population-based study": Supplementary Table 3.pdf

Supplementary Table 3. The quartile distribution of thyroid function/homeostasis parameters in participants with different types of urinary incontinence in the NHANES 2007–2012 cycles.

| Characteristics | Total | SUI | UUI | MUI | Without UI | P-value |
|---|---|---|---|---|---|---|
| FT3 (pg/mL) | | | | | | |
| Q1 | 902 (24.11%) | 206 (24.91%) | 157 (33.25%) | 165 (26.43%) | 374 (20.87%) | 0.025* |
| Q2 | 825 (24.83%) | 223 (26.22%) | 102 (23.16%) | 140 (27.67%) | 360 (23.49%) | |
| Q3 | 1020 (30.62%) | 280 (31.37%) | 93 (28.64%) | 163 (27.15%) | 484 (31.81%) | |
| Q4 | 696 (20.44%) | 163 (17.50%) | 70 (14.95%) | 120 (18.75%) | 343 (23.83%) | |
| FT4 (pmol/L) | | | | | | |
| Q1 | 1223 (29.49%) | 318 (31.28%) | 133 (26.20%) | 213 (28.20%) | 559 (29.68%) | 0.017* |
| Q2 | 1040 (30.70%) | 261 (29.73%) | 127 (29.05%) | 173 (26.86%) | 479 (32.87%) | |
| Q3 | 660 (21.58%) | 172 (23.53%) | 84 (20.01%) | 101 (18.97%) | 303 (21.75%) | |
| Q4 | 520 (18.23%) | 121 (15.47%) | 78 (24.74%) | 101 (25.97%) | 220 (15.69%) | |
| TSH (mIU/L) | | | | | | |
| Q1 | 866 (23.94%) | 202 (19.43%) | 106 (22.64%) | 132 (21.71%) | 426 (27.45%) | 0.043* |
| Q2 | 856 (24.56%) | 207 (25.19%) | 113 (26.13%) | 128 (21.14%) | 408 (25.01%) | |
| Q3 | 861 (27.82%) | 233 (31.79%) | 94 (26.26%) | 153 (28.29%) | 381 (25.85%) | |
| Q4 | 860 (23.68%) | 230 (23.60%) | 109 (24.97%) | 175 (28.87%) | 346 (21.70%) | |
| TG (ng/mL) | | | | | | |
| Q1 | 862 (25.89%) | 221 (25.26%) | 100 (27.68%) | 157 (26.48%) | 384 (25.64%) | 0.045* |
| Q2 | 860 (24.81%) | 235 (29.28%) | 84 (20.08%) | 130 (20.17%) | 411 (24.98%) | |
| Q3 | 860 (24.89%) | 227 (24.52%) | 105 (19.84%) | 145 (25.26%) | 383 (26.09%) | |
| Q4 | 861 (24.41%) | 189 (20.94%) | 133 (32.40%) | 156 (28.09%) | 383 (23.29%) | |
| FT3/FT4 | | | | | | |
| Q1 | 901 (27.71%) | 213 (25.76%) | 145 (36.65%) | 167 (36.24%) | 376 (23.92%) | 0.003* |
| Q2 | 850 (24.94%) | 220 (26.41%) | 103 (22.00%) | 133 (21.43%) | 394 (25.96%) | |
| Q3 | 874 (24.83%) | 229 (25.43%) | 98 (24.03%) | 152 (20.86%) | 395 (26.02%) | |
| Q4 | 818 (22.52%) | 210 (22.40%) | 76 (17.32%) | 136 (21.47%) | 396 (24.10%) | |
| TT4RI | | | | | | |
| Q1 | 861 (23.11%) | 201 (20.22%) | 114 (24.38%) | 127 (18.80%) | 419 (25.85%) | 0.047* |
| Q2 | 861 (25.18%) | 224 (26.11%) | 93 (18.61%) | 135 (24.09%) | 409 (26.50%) | |
| Q3 | 860 (26.51%) | 213 (26.88%) | 106 (31.73%) | 155 (27.23%) | 386 (24.91%) | |
| Q4 | 861 (25.20%) | 234 (26.79%) | 109 (25.28%) | 171 (29.87%) | 347 (22.74%) | |
| TT3RI | | | | | | |
| Q1 | 861 | 195 | 116 | 135 | 415 | 0.047* |

| Characteristics | Total | SUI | UUI | MUI | Without UI | P-value |
|---|---|---|---|---|---|---|
| | (23.74%) | (19.66%) | (25.76%) | (21.78%) | (26.18%) | |
| Q2 | 862 (24.76%) | 215 (24.96%) | 107 (21.59%) | 130 (22.91%) | 410 (25.99%) | |
| Q3 | 859 (26.76%) | 234 (31.67%) | 97 (27.90%) | 144 (24.69%) | 384 (24.52%) | |
| Q4 | 861 (24.73%) | 228 (23.70%) | 102 (24.75%) | 179 (30.62%) | 352 (23.31%) | |
| TFQI_FT4 | | | | | | |
| Q1 | 861 (24.81%) | 213 (24.90%) | 104 (23.33%) | 129 (18.90%) | 415 (27.09%) | 0.220 |
| Q2 | 861 (25.17%) | 213 (23.46%) | 99 (22.41%) | 149 (26.07%) | 400 (26.41%) | |
| Q3 | 861 (24.44%) | 221 (24.78%) | 103 (24.68%) | 151 (26.17%) | 386 (23.62%) | |
| Q4 | 860 (25.58%) | 225 (26.87%) | 116 (29.58%) | 159 (28.86%) | 360 (22.88%) | |
| TFQI_FT3 | | | | | | |
| Q1 | 863 (26.38%) | 202 (25.35%) | 135 (32.95%) | 135 (26.02%) | 391 (25.60%) | 0.647 |
| Q2 | 860 (23.89%) | 204 (24.34%) | 110 (21.76%) | 140 (23.38%) | 406 (24.29%) | |
| Q3 | 859 (23.60%) | 221 (21.68%) | 100 (23.14%) | 155 (25.06%) | 383 (24.26%) | |
| Q4 | 861 (26.13%) | 245 (28.63%) | 77 (22.15%) | 158 (25.54%) | 381 (25.86%) | |
| TSHI | | | | | | |
| Q1 | 861 (22.93%) | 201 (20.22%) | 111 (23.53%) | 124 (18.27%) | 425 (25.83%) | 0.028* |
| Q2 | 863 (25.75%) | 216 (25.49%) | 93 (20.40%) | 144 (25.87%) | 410 (27.05%) | |
| Q3 | 858 (26.41%) | 231 (29.02%) | 105 (29.98%) | 146 (25.88%) | 376 (24.37%) | |
| Q4 | 861 (24.91%) | 224 (25.27%) | 113 (26.09%) | 174 (29.98%) | 350 (22.74%) | |
