## Supplemental Table1-6 for "Association between thyroid function and thyroid homeostasis parameters and female urinary incontinence: A population-based study": Supplementary Table 4.pdf

Supplementary Table 4. Association between thyroid function with urgency urinary incontinence.

| | **Model 1** | | **Model 2** | | **Model 3** | |
|---|---|---|---|---|---|---|
| | OR (95% CI) | P-value | OR (95% CI) | P-value | OR (95% CI) | P-value |
| FT3 (pg/mL) | | | | | | |
| Continuous | 0.61 (0.45–0.83) | 0.003 | 0.86 (0.65–1.14) | 0.293 | 0.91 (0.68–1.22) | 0.539 |
| Q1 | Ref | Ref | Ref | Ref | Ref | Ref |
| Q2 | 0.81 (0.61-1.07) | 0.147 | 0.92 (0.68-1.23) | 0.562 | 1.15 (0.75-1.74) | 0.529 |
| Q3 | 0.67 (0.51-0.87) | 0.004 | 0.87 (0.65-1.16) | 0.359 | 0.97 (0.67-1.42) | 0.885 |
| Q4 | 0.61 (0.43-0.87) | 0.009 | 0.84 (0.58-1.22) | 0.378 | 1.00 (0.65-1.53) | 0.982 |
| p for trend | | 0.002 | | 0.285 | | 0.758 |
| FT4 (pmol/L) | | | | | | |
| Continuous | 1.08 (1.03–1.13) | 0.002 | 1.04 (0.99–1.08) | 0.113 | 1.04 (0.99–1.11) | 0.158 |
| Q1 | Ref | Ref | Ref | Ref | Ref | Ref |
| Q2 | 0.97 (0.73-1.27) | 0.801 | 0.91 (0.68-1.21) | 0.508 | 0.87 (0.58-1.29) | 0.493 |
| Q3 | 0.96 (0.75-1.23) | 0.728 | 0.87 (0.68-1.12) | 0.288 | 0.93 (0.64-1.34) | 0.694 |
| Q4 | 1.80 (1.29-2.51) | 0.001 | 1.41 (0.99-2.01) | 0.065 | 1.62 (1.02-2.59) | 0.054 |
| p for trend | | 0.003 | | 0.105 | | 0.067 |
| TSH (mIU/L) | | | | | | |
| Continuous | 0.99 (0.97–1.01) | 0.468 | 0.98 (0.96–1.01) | 0.246 | 0.97 (0.94–1.01) | 0.120 |
| Q1 | Ref | Ref | Ref | Ref | Ref | Ref |
| Q2 | 1.03 (0.78-1.36) | 0.841 | 0.96 (0.72-1.29) | 0.804 | 0.92 (0.64-1.31) | 0.634 |
| Q3 | 1.10 (0.79-1.53) | 0.592 | 0.97 (0.70-1.36) | 0.879 | 0.98 (0.67-1.42) | 0.913 |
| Q4 | 1.36 (0.99-1.87) | 0.061 | 1.01 (0.71-1.45) | 0.950 | 0.94 (0.59-1.49) | 0.786 |
| p for trend | | 0.051 | | 0.930 | | 0.856 |
| TG (ng/mL) | | | | | | |
| Continuous | 1.00 (1.00–1.01) | 0.060 | 1.00 (1.00–1.01) | 0.115 | 1.00 (1.00–1.01) | 0.184 |
| Q1 | Ref | Ref | Ref | Ref | Ref | Ref |
| Q2 | 0.72 (0.56-0.92) | 0.011 | 0.87 (0.65-1.15) | 0.325 | 0.77 (0.52-1.14) | 0.202 |
| Q3 | 0.86 (0.64-1.15) | 0.303 | 0.93 (0.68-1.28) | 0.672 | 0.96 (0.64-1.44) | 0.836 |
| Q4 | 1.26 (0.90-1.75) | 0.188 | 1.31 (0.93-1.85) | 0.129 | 1.50 (1.00-2.26) | 0.064 |
| p for trend | | 0.143 | | 0.146 | | 0.046 |

Model 1: Non-adjusted.

Model 2: Adjusted for age, race, education level, Marital status, Family poverty ratio and BMI.

Model 3: Adjusted for age, race, education level, Marital status, Family poverty ratio, BMI, Hypertension, Diabetes, Smoking, Drinking, Moderate physical activity, Hugefetus, Hysterectomy, Number of vaginal deliveries and Number of sexual partners.
