## Supplemental Table1-6 for "Association between thyroid function and thyroid homeostasis parameters and female urinary incontinence: A population-based study": Supplementary Table 5.pdf

Supplementary Table 5. Association between thyroid function with mixed urinary incontinence.

| | Model 1 | | Model 2 | | Model 3 | |
|---|---|---|---|---|---|---|
| | OR (95% CI) | P-value | OR (95% CI) | P-value | OR (95% CI) | P-value |
| FT3 (pg/mL) | | | | | | |
| Continuous | 0.79 (0.54–1.16) | 0.241 | 0.97 (0.77–1.22) | 0.796 | 0.98 (0.76–1.26) | 0.876 |
| Q1 | Ref | Ref | Ref | Ref | Ref | Ref |
| Q2 | 1.02 (0.72-1.44) | 0.909 | 1.13 (0.80-1.62) | 0.487 | 1.13 (0.73-1.74) | 0.600 |
| Q3 | 0.78 (0.56-1.08) | 0.141 | 0.95 (0.66-1.37) | 0.793 | 1.03 (0.64-1.66) | 0.893 |
| Q4 | 0.81 (0.49-1.33) | 0.408 | 1.02 (0.61-1.70) | 0.933 | 1.04 (0.60-1.82) | 0.879 |
| p for trend | | 0.170 | | 0.854 | | 0.965 |
| FT4 (pmol/L) | | | | | | |
| Continuous | 1.05 (1.00–1.11) | 0.066 | 1.02 (0.97–1.08) | 0.441 | 1.05 (0.98–1.12) | 0.147 |
| Q1 | Ref | Ref | Ref | Ref | Ref | Ref |
| Q2 | 0.90 (0.66-1.23) | 0.512 | 0.86 (0.65-1.14) | 0.298 | 0.88 (0.62-1.25) | 0.476 |
| Q3 | 0.91 (0.67-1.23) | 0.532 | 0.85 (0.61-1.18) | 0.337 | 0.88 (0.56-1.39) | 0.587 |
| Q4 | 1.63 (1.12-2.37) | 0.013 | 1.35 (0.89-2.05) | 0.161 | 1.80 (1.08-2.99) | 0.035 |
| p for trend | | 0.029 | | 0.229 | | 0.051 |
| TSH (mIU/L) | | | | | | |
| Continuous | 1.00 (0.98–1.02) | 0.990 | 0.99 (0.97–1.02) | 0.659 | 0.97 (0.94–1.01) | 0.137 |
| Q1 | Ref | Ref | Ref | Ref | Ref | Ref |
| Q2 | 0.94 (0.64-1.38) | 0.756 | 0.88 (0.60-1.30) | 0.535 | 0.97 (0.58-1.64) | 0.916 |
| Q3 | 1.15 (0.79-1.66) | 0.481 | 1.00 (0.68-1.49) | 0.981 | 1.04 (0.66-1.65) | 0.858 |
| Q4 | 1.43 (1.00-2.03) | 0.054 | 1.06 (0.72-1.57) | 0.760 | 0.94 (0.55-1.60) | 0.810 |
| p for trend | | 0.035 | | 0.607 | | 0.890 |
| TG (ng/mL) | | | | | | |
| Continuous | 1.00 (1.00–1.00) | 0.141 | 1.00 (1.00–1.00) | 0.219 | 1.00 (1.00–1.00) | 0.332 |
| Q1 | Ref | Ref | Ref | Ref | Ref | Ref |
| Q2 | 0.76 (0.59-0.99) | 0.052 | 0.89 (0.66-1.20) | 0.450 | 0.88 (0.61-1.28) | 0.513 |
| Q3 | 0.99 (0.68-1.45) | 0.962 | 1.07 (0.74-1.54) | 0.718 | 1.06 (0.65-1.74) | 0.805 |
| Q4 | 1.15 (0.79-1.68) | 0.462 | 1.22 (0.84-1.76) | 0.304 | 1.47 (0.89-2.43) | 0.148 |
| p for trend | | 0.341 | | 0.278 | | 0.138 |

Model 1: Non-adjusted.

Model 2: Adjusted for age, race, education level, Marital status, Family poverty ratio and BMI.

Model 3: Adjusted for age, race, education level, Marital status, Family poverty ratio, BMI, Hypertension, Diabetes, Smoking, Drinking, Moderate physical activity, Hugefetus, Hysterectomy, Number of vaginal deliveries and Number of sexual partners.
