## Supplemental Table1-6 for "Association between thyroid function and thyroid homeostasis parameters and female urinary incontinence: A population-based study": Supplementary Table 6.pdf

Supplementary Table 6. Association between thyroid homeostasis parameters with stress urinary incontinence.

| | **Model 1** | | **Model 2** | | **Model 3** | |
|---|---|---|---|---|---|---|
| | OR (95% CI) | P-value | OR (95% CI) | P-value | OR (95% CI) | P-value |
| FT3/FT4 | | | | | | |
| Continuous | 1.26 (0.59–2.69) | 0.550 | 1.85 (0.52–6.58) | 0.348 | 1.05 (0.50–2.23) | 0.892 |
| Q1 | Ref | Ref | Ref | Ref | Ref | Ref |
| Q2 | 1.19 (0.91-1.56) | 0.201 | 1.34 (1.01-1.78) | 0.047 | 1.55 (1.12-2.14) | 0.016 |
| Q3 | 1.14 (0.87-1.50) | 0.359 | 1.31 (0.94-1.83) | 0.114 | 1.31 (0.93-1.86) | 0.138 |
| Q4 | 1.09 (0.85-1.41) | 0.484 | 1.26 (0.96-1.65) | 0.107 | 1.22 (0.86-1.73) | 0.285 |
| p for trend | | 0.493 | | 0.108 | | 0.422 |
| TT4RI | | | | | | |
| Continuous | 1.00 (0.99–1.00) | 0.365 | 1.00 (0.99–1.00) | 0.174 | 1.00 (0.99–1.00) | 0.210 |
| Q1 | Ref | Ref | Ref | Ref | Ref | Ref |
| Q2 | 1.25 (0.93-1.69) | 0.145 | 1.19 (0.90-1.59) | 0.225 | 0.94 (0.65-1.36) | 0.742 |
| Q3 | 1.22 (0.90-1.63) | 0.202 | 1.08 (0.79-1.47) | 0.622 | 1.01 (0.71-1.44) | 0.966 |
| Q4 | 1.30 (0.93-1.80) | 0.129 | 1.08 (0.78-1.51) | 0.630 | 1.00 (0.69-1.44) | 0.979 |
| p for trend | | 0.134 | | 0.826 | | 0.906 |
| TT3RI | | | | | | |
| Continuous | 1.00 (0.99–1.00) | 0.279 | 0.99 (0.98–1.00) | 0.244 | 0.99 (0.99–1.00) | 0.110 |
| Q1 | Ref | Ref | Ref | Ref | Ref | Ref |
| Q2 | 1.29 (0.98-1.71) | 0.075 | 1.26 (0.96-1.65) | 0.104 | 1.26 (0.92-1.73) | 0.168 |
| Q3 | 1.62 (1.25-2.09) | 0.001 | 1.49 (1.13-1.95) | 0.007 | 1.31 (0.92-1.86) | 0.144 |
| Q4 | 1.21 (0.90-1.63) | 0.214 | 1.04 (0.78-1.39) | 0.798 | 1.11 (0.82-1.52) | 0.501 |
| p for trend | | 0.084 | | 0.587 | | 0.501 |
| TFQI_FT4 | | | | | | |
| Continuous | 1.07 (0.86–1.34) | 0.541 | 0.89 (0.69–1.14) | 0.350 | 1.04 (0.76–1.42) | 0.808 |
| Q1 | Ref | Ref | Ref | Ref | Ref | Ref |
| Q2 | 0.91 (0.66-1.24) | 0.543 | 0.85 (0.62-1.18) | 0.342 | 0.84 (0.55-1.29) | 0.441 |
| Q3 | 1.01 (0.75-1.38) | 0.928 | 0.92 (0.67-1.26) | 0.609 | 1.02 (0.70-1.50) | 0.910 |
| Q4 | 1.06 (0.81-1.41) | 0.662 | 0.89 (0.66-1.20) | 0.464 | 1.01 (0.71-1.43) | 0.971 |
| p for trend | | 0.499 | | 0.599 | | 0.709 |
| TFQI_FT3 | | | | | | |
| Continuous | 1.07 (0.82–1.40) | 0.614 | 1.05 (0.81–1.37) | 0.705 | 1.16 (0.86–1.56) | 0.329 |
| Q1 | Ref | Ref | Ref | Ref | Ref | Ref |
| Q2 | 1.08 (0.79-1.49) | 0.628 | 1.13 (0.82-1.54) | 0.460 | 1.07 (0.70-1.62) | 0.763 |
| Q3 | 0.94 (0.68-1.30) | 0.721 | 0.95 (0.70-1.29) | 0.759 | 1.03 (0.69-1.53) | 0.891 |
| Q4 | 1.20 (0.90-1.59) | 0.229 | 1.19 (0.89-1.59) | 0.259 | 1.19 (0.86-1.65) | 0.317 |
| p for trend | | 0.356 | | 0.427 | | 0.375 |
| TSHI | | | | | | |
| Continuous | 1.09 (0.98–1.21) | 0.139 | 1.01 (0.89–1.15) | 0.860 | 1.06 (0.92–1.23) | 0.435 |
| Q1 | Ref | Ref | Ref | Ref | Ref | Ref |
| Q2 | 1.16 (0.87-1.55) | 0.308 | 1.10 (0.83-1.46) | 0.509 | 0.92 (0.62-1.36) | 0.688 |
| Q3 | 1.34 (1.02-1.76) | 0.039 | 1.18 (0.89-1.57) | 0.266 | 1.17 (0.84-1.65) | 0.362 |
| Q4 | 1.20 (0.88-1.64) | 0.244 | 1.00 (0.73-1.35) | 0.975 | 0.96 (0.66-1.38) | 0.809 |
| p for trend | | 0.140 | | 0.946 | | 0.781 |

Model 1: Non-adjusted.

Model 2: Adjusted for age, race, education level, Marital status, Family poverty ratio and BMI.
Model 3: Adjusted for age, race, education level, Marital status, Family poverty ratio, BMI, Hypertension, Diabetes, Smoking, Drinking, Moderate physical activity, Hugefetus, Hysterectomy, Number of vaginal deliveries and Number of sexual partners.
